# COVID-19 healthcare demand projections: Arizona

**DOI:** 10.1101/2020.05.13.20099838

**Authors:** Esma S. Gel, Megan Jehn, Timothy Lant, Anna R. Muldoon, Trisalyn Nelson, Heather M. Ross

## Abstract

Beginning in March 2020, the United States emerged as the global epicenter for COVID-19 cases. In the ensuing weeks, American jurisdictions attempted to manage disease spread on a regional basis using non-pharmaceutical interventions (i.e., social distancing), as uneven disease burden across the expansive geography of the United States exerted different implications for policy management in different regions. While Arizona policymakers relied initially on state-by-state national modeling projections from different groups outside of the state, we sought to create a state-specific model using a mathematical framework that ties disease surveillance with the future burden on Arizona’s healthcare system. Our framework uses a compartmental system dynamics model using a SEIRD framework that accounts for multiple types of disease manifestations for the COVID-19 infection, as well as the observed time delay in epidemiological findings following public policy enactments. We use a bin initialization logic coupled with a fitting technique to construct projections for key metrics to guide public health policy, including exposures, infections, hospitalizations, and deaths under a variety of social reopening scenarios.

## Introduction

Since its documented onset in December 2019 and formal identification in January 2020 in Wuhan, China, COVID-19 (SARS-CoV-2) has spread around the globe, infecting more than 7.5 million people globally by mid June 2020 [5]. In an atmosphere of intense uncertainty around many of the epidemiological parameters for modeling including true case counts as a result of low testing availability, the Modeling Emerging Threats for Arizona (METAz) Workgroup of Arizona State University has developed and refined models for predicting the burden of disease in order to inform policy related to nonpharmaceutical interventions (i.e., social distancing). Because the burden of disease and transmission dynamics differ by location due to a variety of factors including geography, population, and environmental conditions, METAz chose to focus on state-level modeling to inform public health response efforts with greater precision. The modeling approaches we describe can be applied to any region or state where region-specific data are available. Here, we focus on the state of Arizona in the American Southwest (population of around 7.3 million, 113,990 sq. miles, majority population concentrated in centrally-located Maricopa County) as a proof of concept.

Arizona’s governor declared a state of emergency on March 11, and municipal governments began to enact limits on in-person gatherings and some business closures on March 16-17. From March 30 to May 15, Arizona was under a stay-at-home order issued by the governor. As of June 10, Arizona has reported 29,852 cases across the state, with the majority of cases in centrally-located Maricopa County, which includes 60 percent of the state’s population. Twenty-seven percent of the total cases were recorded in the first ten days of June. As of June 10, Arizona’s healthcare system has not experienced an overwhelming surge of COVID-19 cases exceeding systemwide capacity to care for critically ill patients. Hospital admissions appeared to have slowed and plateaued in April and May, indicating that social distancing motivated by state and municipal policies enacted beginning in mid-March had reduced transmission and may have been flattening the curve effectively in order to allow time to prepare operations for future management of the disease in Arizona and avoid overwhelming hospital systems as other states experienced.

However, as of early June, Arizona is experiencing increasing widespread community transmission of SARS-CoV-2. Due to a relatively low rate of testing statewide, there is ongoing debate and uncertainty about whether Arizona’s case prevalence data provides an accurate portrait of the true public health risk burden and whether we have passed an (initial) peak of infections and hospitalizations statewide and in individual counties. Projections from a variety of modeling groups (i.e., IHME, UA, ASU) had indicated that the peak number of cases will be reached in Arizona in mid-April to mid-May. However, it is important to note that modeling projections are inherently uncertain, and accurate assessment of case peaks will be possible only once the peak has passed. In light of the transmission dynamics and laboratory reporting delays for the SARS-CoV-2 outbreak, peak determination will be possible approximately two to four weeks following peak occurrence. It is also important to note that there is still significant uncertainty about the transmission dynamics of the virus, including the degree of asymptomatic infection and transmission and the results do not capture the full range of uncertainty. We demonstrate this observation through our modeling below.

On April 16, the United States Government released Guidelines for Opening Up America Again, proposing a phased approach to reopening the country. In order to progress into and through three sequential phases of opening businesses and other public and private services, states are expected to meet a set of gating criteria outcome metrics along with a set of capacity responsibilities for carrying out core public health and management functions. In order to move to Phase 1 with limited reopening of businesses and other services, states must demonstrate flattening the case rates, and in order to move to Phase 2 with expanded reopening of businesses and services, states must demonstrate no rebound in case counts from the limited reopening in Phase 1.

On May 15, Arizona’s stay-at-home order expired, with targeted business openings occurring on May 8 and May 11. At the time of reopening, Arizona had not met the CDC gating criteria to move to Phase 1, nor had the state developed a comprehensive plan that incorporated the full testing capabilities within the state (both molecular and serological) with a program linked to non-pharmaceutical interventions (NPI) including stay-at-home and other social distancing and infection mitigation policies and procedures. In order to reopen Arizona safely, a phased approach needs to be data-driven and focused on avoiding a rapid surge in cases through appropriate and effective policy for non-pharmaceutical interventions.

Rising case counts and hospitalizations in late May and early June reflect that the move to lift policies restricting in-person interactions and the lack of statewide policies to enforce NPIs including physical distancing, masking, and hand hygiene resulted in markedly increased community transmission. As of June 10, there is not a statewide plan articulated to guide resumption of NPIs despite strong evidence of increased community transmission.

This paper proposes a mathematical framework that ties disease surveillance with the future burden on Arizona’s hospital system and hospital resources. The mathematical model links together policy interventions with estimated outcomes for infections, hospitalizations, and deaths in an epidemiological analysis. One of the key features of our modeling methodology is the time-delay of new infections on confirmed case counts and the impact on the healthcare system. We propose methods to evaluate the likely outcomes for a range of policy decisions intended to keep Arizona safe while reopening in a responsible and defensible sequence.

## 1 Methods

### 1.1 Data Sources

We use two publicly available data sources to initialize and fit our model: cumulative case counts and deaths in the State of Arizona between the dates of March 4 and June 7. Figures 1 and 2 depict the data that are used to obtain the results presented below. Both of these are publicly available and daily announced at the Arizona Department of Health Services’ (ADHS) data dashboard at https://www.azdhs.gov/preparedness/epidemiology-disease-control/infectious-disease-epidemiology/covid-19/dashboards/index.php.

**Fig 1.**
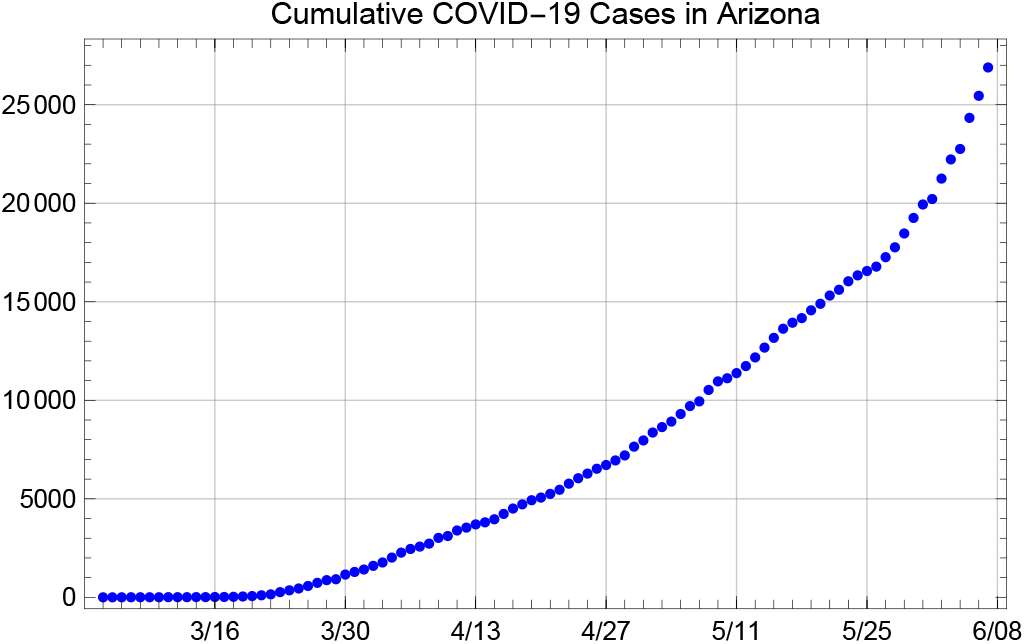
Cumulative confirmed COVID-19 cases in Arizona, between March 4 to June 7, 2020

**Fig 2.**
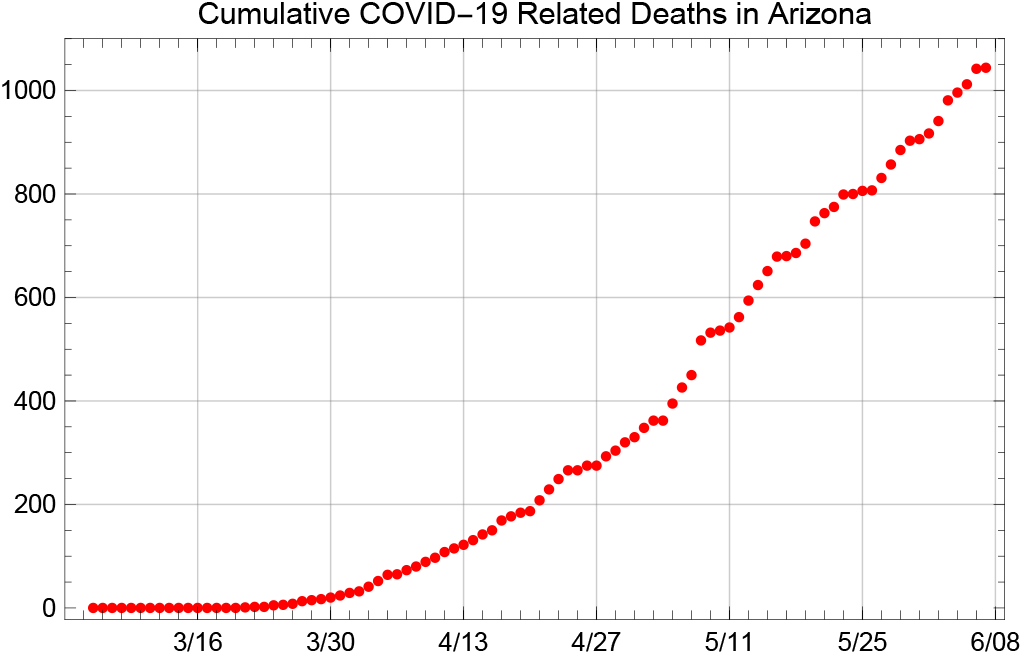
Cumulative COVID-19 related deaths in Arizona, between March 4 to June 7, 2020

### 1.2 Structure of the Model

We make use of a compartmental system dynamics model using a SEIRD framework that includes multiple compartments for infected individuals. This model structure allows us to estimate the number of patients in the hospital and assess model fit with respect to two sources of data: daily new cases obtained from the daily cumulative confirmed cases and daily cumulative reported deaths given in Figures 1 and 2. In essence, the population of interest, in this case, the population of the State of Arizona (assumed to be 7,278,717 in this study) is divided into states of Susceptible 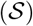, Exposed but not yet infectious 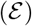, Asymptomatic infected 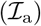, infectious and presymptomatic 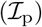, Symptomatic with a mild infection 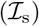, symptomatic with a severe infection and hospitalized 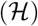, symptomatic with a critical infection and in the ICU 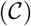, undergoing additional recovery in ICU 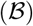, Recovered and immune 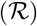 and Dead 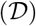, as shown in Figure 3.

**Fig 3.**
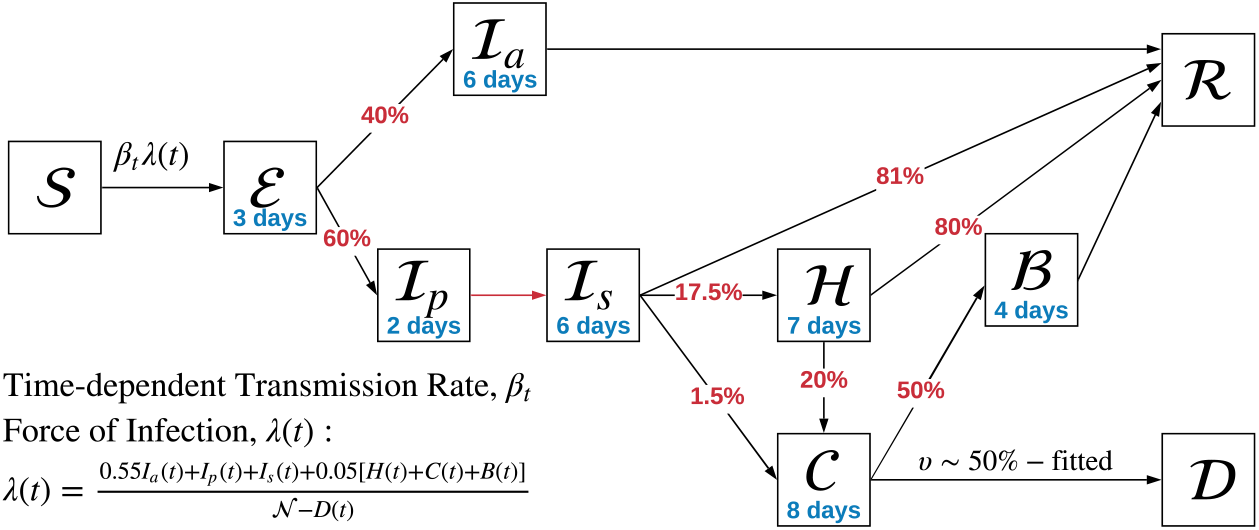
Depiction of the compartmentalized system dynamics model used to represent transmission and disease progression for State of Arizona projections

Our model defines separate bins for asymptomatic and presymptomatic individuals to explicitly account for differing rates of transmission and differing durations of viral shedding. Individuals who are exposed to the virus generally go through an incubation period (modeled by a rate of *ζ*) during which they are exposed but not yet infectious. This duration is modeled as 3 days in our study, to support an ensemble estimate of 5 days for time from exposure to symptoms and an estimate of 6 days serial time obtained from the literature [1, 6, 10, 13, 14, 23]. After the preinfectious period, individuals become infectious, either as an asymptomatic or a presymptomatic patient. The presymptomatic duration (modeled with rate *δ*) is assumed to be 2 days [20, 24]. Asymptomatic patients recover at a rate of *γ* = 1/6, corresponding to an average recovery duration of 6 days after the preinfectious period. We modeled a number of observed variations on how symptomatic individuals experience COVID-19. After the presymptomatic period of 2 days, a large fraction, 81% (denoted by *ρ* in the model) estimated by [22, 25], of individuals go through a 6-day period of relatively mild symptoms and recover similarly to the asymptomatic patients [21]. The remaining 19% of symptomatic patients develop a severe or critical infection and seek care at a hospital.

According to the growing body of peer-reviewed literature, a large portion of the patients admitted to the hospital have a severe, but not critical, infection and recover after an average duration of 7 days in a regular hospital bed. An average of 20% of these patients, however, progress to a critical infection, requiring ICU care and possibly intubation. In addition to patients that progress to the ICU from a regular hospital bed, a small fraction of patients that present to the hospital with critical respiratory distress are directly admitted to the ICU. We have conferred with local clinicians in the Phoenix metropolitan area and Tucson who confirm these patterns of patient progression through the hospital system. Hence, in our model, there are two modes of admission to the ICU; one directly from the emergency room and the other one from a regular ward, after the patient’s infection progresses to a critical condition. The parameters for these splits are set to ensure that (i) the fraction of symptomatic patients with mild infection is 81% [22, 25], (ii) the total fraction of symptomatic patients that develop a critical infection that requires ICU care is 5% [22] and (iii) 20% of patients in a regular bed progress to a critical infection [16].

Studies in the literature cite a diverse range of outcomes for patients in the ICU, but most agree that the ICU duration for patients that eventually recover is generally longer. For example, one study [19] cites point estimates for the duration of onset-of-symptoms to death to be 17.8 days and from onset-of-symptoms to hospital discharge to be 22.6 days. The additional time to discharge is due partly to various steps that caregivers have to take to arrange for care after the ICU period since generally patients that underwent intubation and other invasive procedures require subsequent care in other post-acute facilities. The additional post-acute recovery time is represented as another bin, with a duration of 4 days (modeled with rate *α*). The reported average ICU stays in the literature are generally very diverse; we adopted a conservative point estimate of 8 days to align with the symptom onset to recovery/death estimates [19] as well as other more detailed studies that tracked patients’ progress through the hospital [25].

One of the important parameters in the model is *ω*, which represents the fraction of asymptomatic patients. Several studies point to the importance of modeling transmissions by asymptomatic individuals, who may never be aware that they were transmitting the virus. However, point estimates on the fraction of individuals that experience asymptomatic infection vary greatly from context to context.

In our models we adopted an asymptomatic rate of 40% based on point estimates observed in multiple peer-reviewed manuscripts from different COVID-19 populations around the world [3, 17]. This assumption allows us to obtain worst-case estimates on the prevalence of infections in the general population given that, in the absence of widespread testing of asymptomatic individuals, the asymptomatic patients are generally undetected.

In our modeling and analysis, we explicitly consider the possibility that only a small fraction of the true incidence of infections are detected as COVID-19 cases and reflected in the reported case counts and deaths. One such example that points to a large undetected fraction of cases is [11], indicating that 86% of the early infections in China were undocumented, or in other words the “actual” cases in a population may be more than 7 times the detected cases. The same study also offers a rate of transmissions by asymptomatic individuals at 55% of the transmission rate by symptomatic individuals, which we reflect in the force of infection, *λ*(t) shown in Figure 3. Subsequently several other papers have offered additional understanding on the role of asymptomatic infections in transmission and its prevalence in different contexts [2, 3, 8, 9, 15, 17]. We use these papers along with the actual data on new cases and deaths in Arizona to obtain point estimates for model parameters. We also devise an initialization algorithm to identify initial values of the compartments in the model.

The ordinary differential equations (ODE) that define the system dynamics are given by Eqns (1) thru (10).

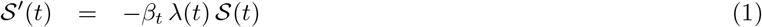

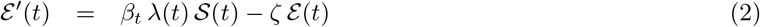

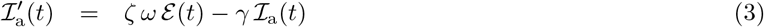

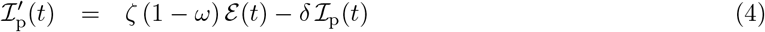

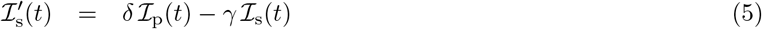

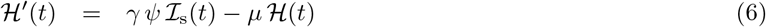

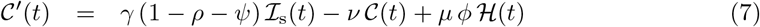

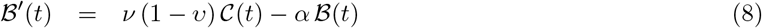

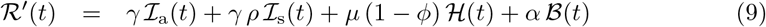

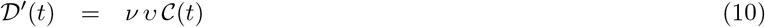

The time dependent force of infection, *λ*(t) is modeled as

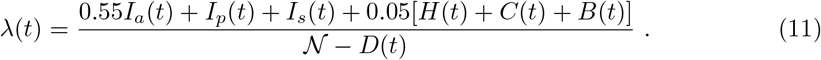

This expression is motivated by the fact that asymptomatic individuals transmit the disease at a reduced rate as discussed above, and patients under care in the hospital are relatively well isolated via institutional infection control measures so they only transmit at a rate that is equal to 5% of the presymptomatic or symptomatic patients. Studies that point to the high infectiousness of presymptomatic patients [4] imply that infections are mostly driven by patients in these compartments. We model a time-dependent transmission rate, *β_t_*, denoted by the subscript *t* to represent the time dependency. Together with the force of infection and the current pool of susceptible individuals, the transmission rate *β_t_* yields the rate at which susceptible individuals get exposed to the infection. The force of infection term can be thought of as the probability that an arbitrary individual is infectious at a rate equivalent to that of a symptomatic patient.

The transmission rate, *β_t_* represents the average rate of contact between susceptible and (symptomatic-equivalent) infectious people multiplied by the probability of transmission given contact. Hence, given the above force of infection and the number of individuals in the susceptible compartment, the rate at which individuals become exposed to the virus at time *t* is strongly driven by the term, *β_t_*. A good way of thinking about the impact of non-pharmaceutical interventions such as social distancing, stay-at-home orders, school closures, wearing masks, etc. is through the term *β_t_*, and how the different interventions impact either (i) the average number of infectious individuals that susceptible individuals contact, or (ii) the probability of transmission given contact.

Note that an increase in either of these two values would lead to an increase in the effective transmission rate at a given time, which will then increase the rate at which susceptible individuals get exposed to the virus. We find Figure 4 to be informative to understand the effect of these two variables to understand the impact of social distancing and other NPI interventions. Staying on the same *β* curve of 0.20 while increasing the average number of contacts for a susceptible individual from 10 to, say, 20 requires that the probability of transmission given contact be reduced from 0.02 to 0.013 through measures that reduce the probability of transmission given contact with an infectious individual. Such measures may involve hand washing practices, wearing masks, keeping 6+ ft apart, etc. As the interactions between individuals are expected to increase after the stay-at-home orders are lifted, the importance of such measures should be more rigorously emphasized. It is also useful to note that a modest 15% increase in both values would result in a 32.25% increase in *β*, which would have dire consequences for the transmission dynamics.

**Fig 4.**
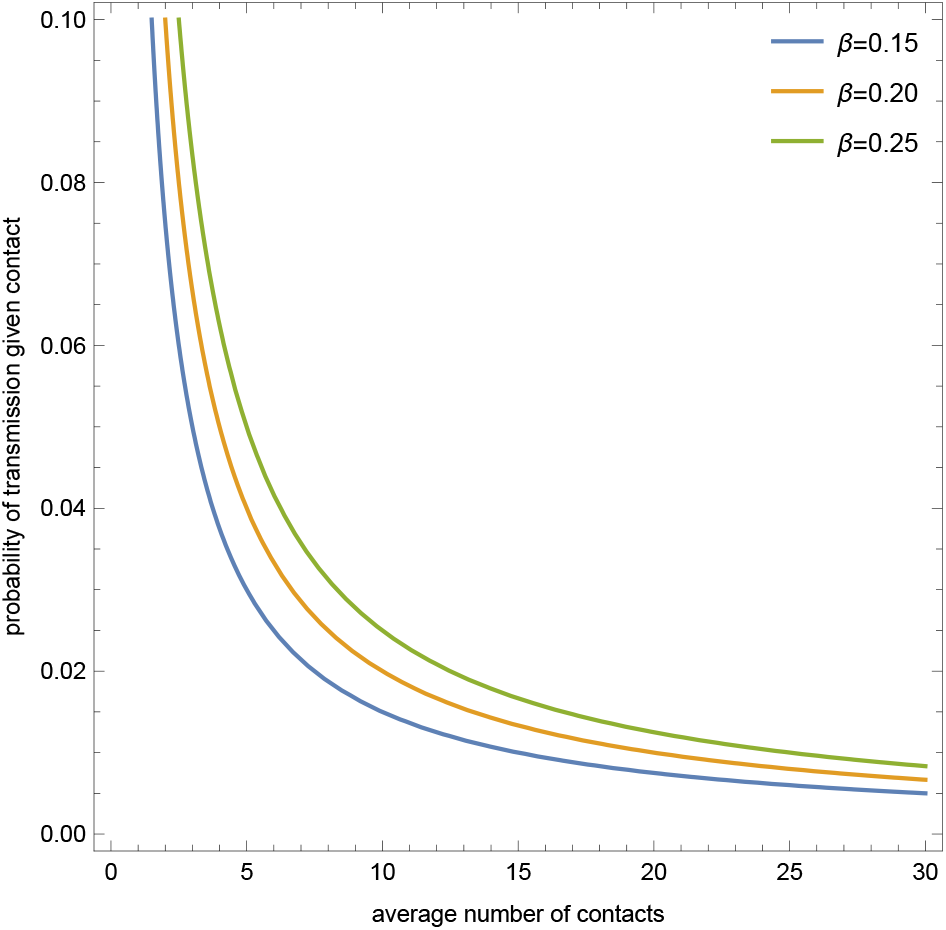
Transmission rate, *β* represented as a product of the average number of contacts and the probability of transmission given contact with an infectious individual

The model parameters and point estimates for them obtained from the literature are provided in Table 1. Our approach of initializing the compartments and fitting the transmission rate *β_t_* and the mortality rate at the ICU, *υ*, uses publicly available data on case counts and COVID-19 related deaths in Arizona. In our fitting procedure, we allow for the transmission rate, *β_t_* to change in response to significant events or policy changes, such as non-pharmaceutical interventions being enacted or lifted in Arizona, as we explain below.

### 1.3 Initialization of Compartments

We first present a methodology to initialize the model in a manner that is independent of the transmission rate, *β*. In particular, we consider the data on the cumulative number of confirmed cases in Arizona, where the first reported cases of community transmission were on March 4, as shown in Figure 1. We use these data to obtain the number of new cases on each day. The average reporting delay on COVID-19 tests is about 6 days in Arizona. Given that our model indicates an incubation period of 5 days and average time to seek testing (when it is available) is about 3 days after symptom-onset, we obtain presumed exposure dates for the reported new cases on each day (i.e., 14 days before a case is confirmed). A visual that shows this logic is shown with the blue bars (reported new cases over time) and the orange bars (numbers eventually detected, shown on the presumed exposure dates) in Figure 5. Note that the orange bars show the number of individuals exposed to the virus on the given day, who are then eventually detected by testing.

As discussed above, a large portion of the individuals exposed to the virus on a given day are never detected due to the fact that (i) a significant portion of these individuals never develop symptoms; and (ii) some symptomatic individuals are never tested, their infections are attributed to another influenza-like illness, or their case is missed due to false negative results in COVID-19 tests. To account for the large rate of undetected infections, we have devised a novel approach using an X-factor initialization scheme where we multiply the number of eventually detected-exposed individuals by the X-factor to obtain the underlying overall exposures on a given presumed exposure day. The X-factor determination in this scheme is highly correlated to the degree to which the testing procedures are able to detect the infections in the system. Given our assumption that 40% of all infections are asymptomatic, the minimum X-factor that is aligned with our modeling assumptions is 1.67, since these individuals are almost never tested and confirmed due to the fact that they do not exhibit symptoms to prompt testing. At the upper end, our model indicates that about 12% of infections have severe or critical infections, requiring them to seek healthcare. This implies that the maximum X-factor that would be aligned with our model is about 8, since nearly all individuals seeking care in Arizona for COVID-like symptoms are tested for COVID-19. In Figure 5, the grey points depict exposures in a scenario using X-factor of 4.

**Table 1.** Point estimates used for model parameters and sources

| Description | Parameter | Value | Sources |
| --- | --- | --- | --- |
| Time to infectiousness | $\zeta^{-1}$ | 3 days | [1, 7, 10, 12, 13, 18, 23] |
| Presymptomatic duration | $\delta^{-1}$ | 2 days | [20, 24] |
| Asymptomatic infectious period | $\gamma^{-1}$ | 6 days | [21, 22] |
| Mild infection recovery time | $\gamma^{-1}$ | 6 days | [22] |
| Severe infection recovery time | $\mu^{-1}$ | 7 days | [6, 22] |
| Critical infection to death | $\nu^{-1}$ | 8 days | [19, 25] |
| Additional days to recover after ICU | $\alpha^{-1}$ | 4 days | [19] |
| Fraction of asymptomatic infections | $\omega$ | 40% | [3, 17] |
| Fraction of mild symptomatic infections | $\rho$ | 81% | [22] |
| Fraction hospitalized on regular bed | $\psi$ | 17.5% | [22] |
| Fraction of hospitalized progressing to ICU | $\phi$ | 20% | [16] |
| Mortality among ICU patients | $v$ | 50-60% | data fit |

**Fig 5.**
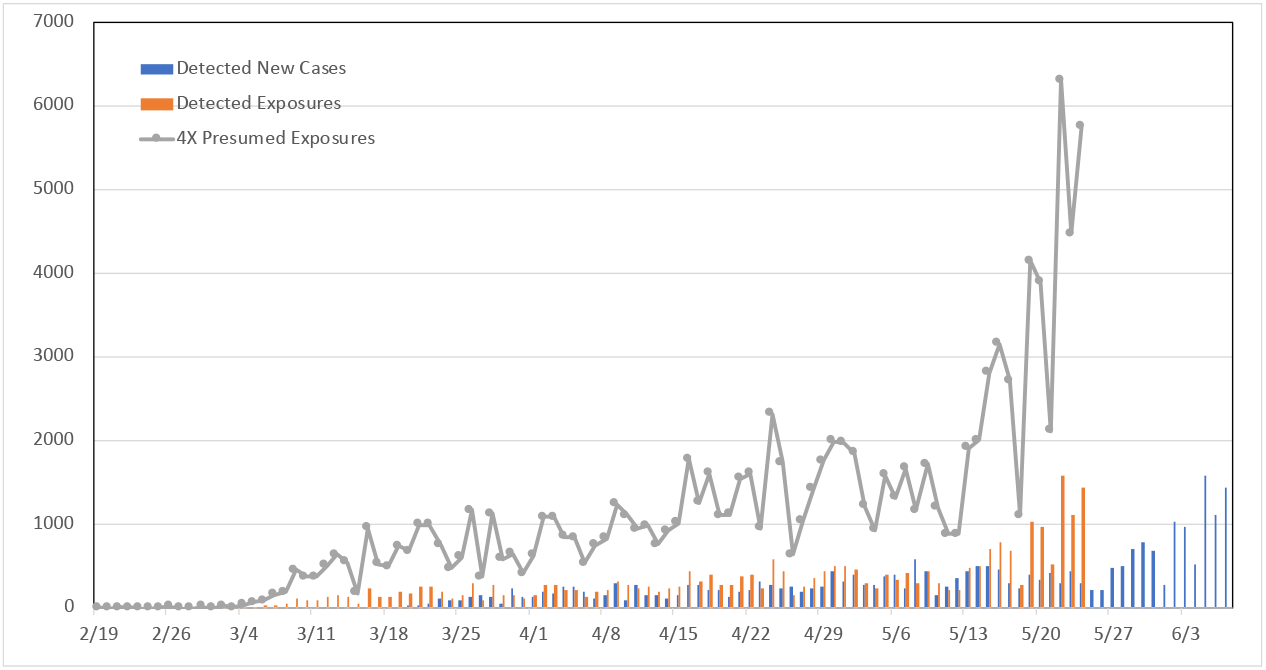
Reported new cases and presumed exposure dates

The X-factored exposures on presumed exposure days are then fed into our SEIRD model, keeping the transmission rate to zero. We obtain an approximate continuous time function by interpolating over these presumed exposures, called *W*(*s*). Figure 6 shows the approximated rate of exposures over time in X-factor of 4 scenario between March 4 and March 29 in Arizona; the black dots are the daily presumed exposures also shown in Figure 5.

**Fig 6.**
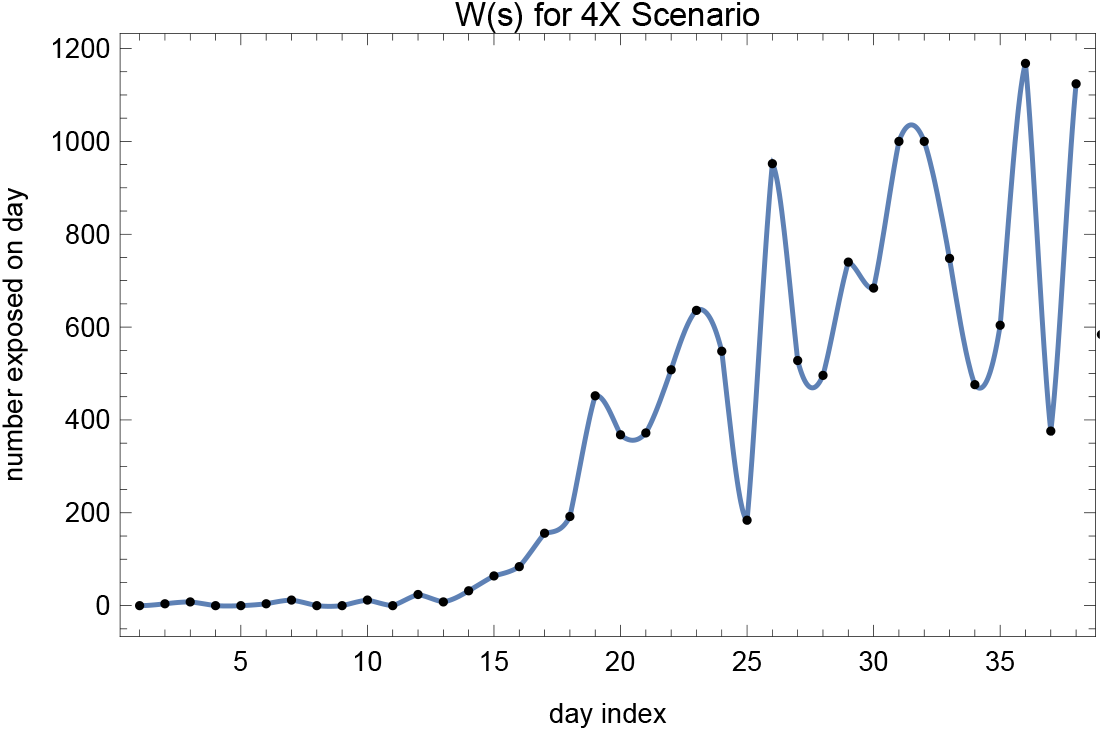
The *W*(*s*) function for the X-factor of 4 scenario, obtained by inflating the daily new cases

We then numerically evaluate the convolution

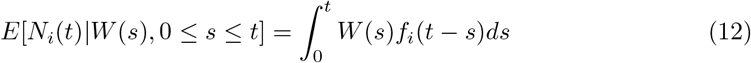

to obtain the expected number in compartment 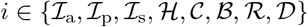 at time *t*, where *f_i_*(*τ*) denotes the probability that an individual would be in Bin i τ time units after exposure to the virus. The *f_i_*(·) functions for each bin in the model can be obtained by simulating the above stated model with one exposed individual and transmission rate of zero. As an example, Figure 7 shows the fraction at the hospital, 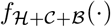 versus time.

**Fig 7.**
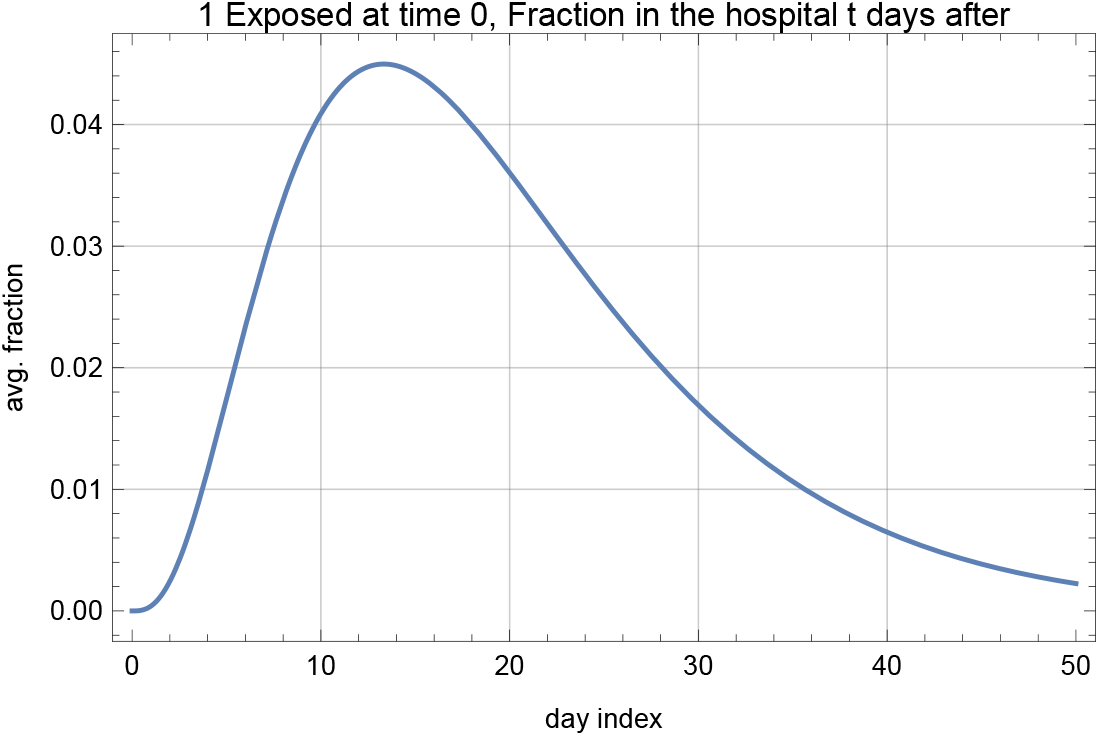
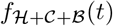 under assumed parameters

The solution to the ODEs is unique given a set of initial values for the number in each bin at time zero. Using the above initialization logic, we calculate the number that we expect to see in each bin on a chosen presumed exposure day, using all of the data on new cases reported on the presumed exposure days prior to this point, and using the number of presumed exposures on that day to initialize the 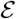 compartment. We are then -almost-ready to simulate the model starting from that day and observe the number in each compartment to obtain projections.

### 1.4 Fitting Transmission Rate and Mortality

In our study, we initialize the bins on March 30 (i.e., this calendar day is our *t* = 0) and use the actual data on presumed exposures (under any assumed X-factor scenario) starting from March 31 until May 24 to fit the transmission rate, *β_t_*. Note that the presumed exposure date of May 24 corresponds to the actual reporting date of June 7, which is the last data point that we use in our results in this manuscript. We tried a number of different initialization dates, and the results on the transmission rate fit were comparable. We divided the time between March 31 and May 24 for which we have presumed exposures data into three periods correlating to dates of significant changes in Arizona public policy and activities related to NPIs including business closures and stay-at-home orders. We fitted three possibly different *β* values to each period, resulting in a piece-wise constant transmission rate structure. In particular, we assumed constant *β* values between March 31 to April 15 (which we refer to as 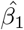, representing early adjustment to the stay-at-home order enacted on March 30), April 16 to May 10 (which we refer to as 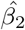, representing stabilization of public response to the stay-at-home order) and May 11 through May 24 (which we refer to as 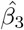, representing reopening of some businesses and activities including personal care services on May 8, dine-in restaurants on May 11, and the expiration of the stay-at-home order on May 15). We assume that 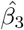 is the best available transmission rate estimate explaining the exposures beyond May 24 (since at the writing of this manuscript no changes in the non-pharmaceurical interventions have been announced) and use that value to generate the projections below for exposure dates later than May 24.

We use Wolfram Mathematica 12 to obtain a numerical solution to the ODEs and obtain a parametric function that describes the number of susceptibles in the system given the initial population of 7,278,717 (population of Arizona) and the assumed loading scenario. We then use the X-factored presumed exposures to obtain the number of susceptibles indicated by the data, and use a least-squares based nonlinear model fit procedure to estimate 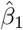, 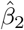 and 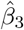. As an example, Figure 8 shows the model fit along with the 95% prediction intervals under a 4X scenario. In addition, we plot the model predicted and 4X presumed exposures in Figure 9.

**Fig 8.**
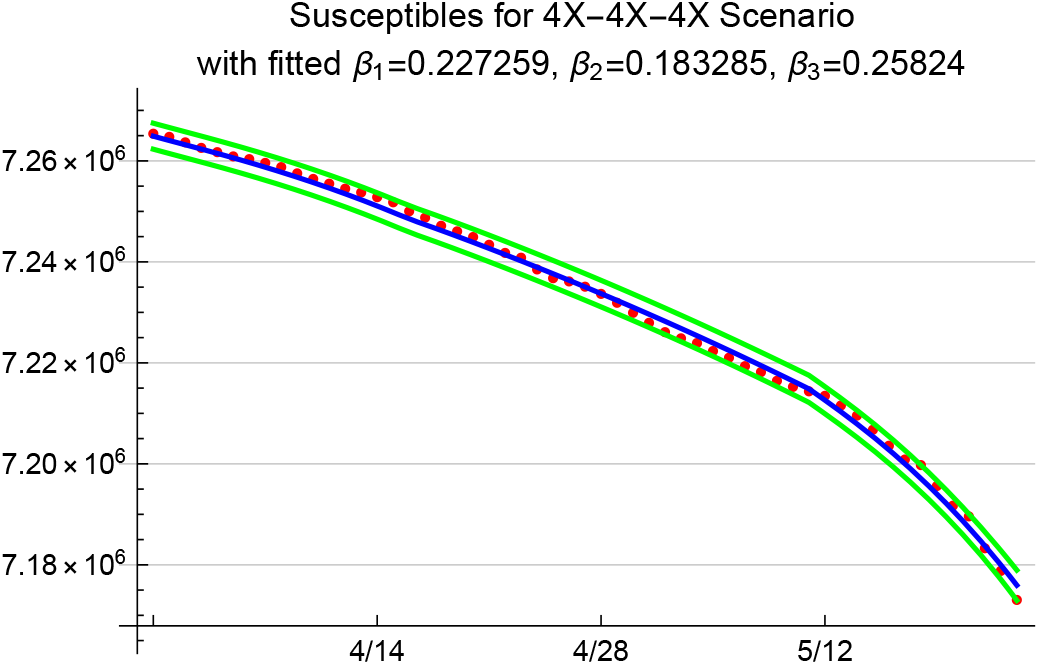
95% prediction bands for susceptibles; red dots show presumed susceptibles under 4X scenario

In addition to fitting the transmission rate, *β*, we use the cumulative number of COVID-19 related deaths in Arizona to fit the mortality rate among the ICU patients. Note that in our model, we assume that all patients who die will do so in the ICU, which ignores the deaths that occur outside the hospital. At the time of the writing of this manuscript, Arizona’s healthcare capacity, beds, and ICU have been sufficient to care for COVID-19 patients. Therefore, to our knowledge, Arizona has not experienced significant reported deaths outside the hospitals due to an inability for patients to access critical care services. There is, however, ongoing debate about whether COVID-19 related deaths are under-reported in Arizona and nationwide.

**Fig 9.**
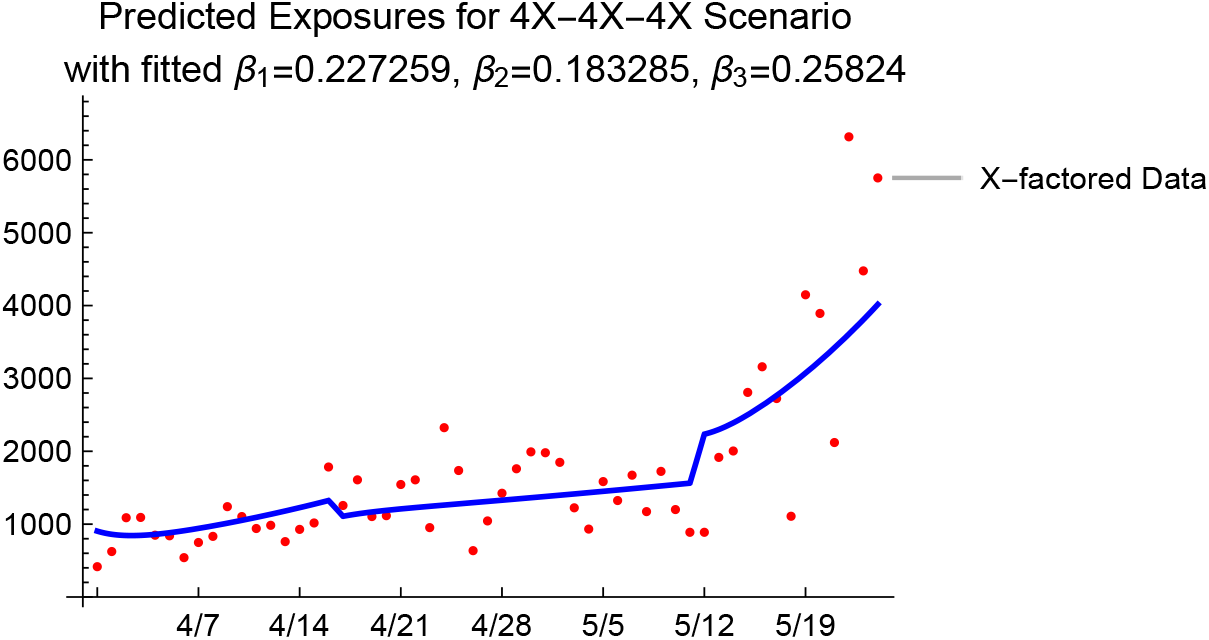
4X presumed exposures from data, and predicted exposures with the fitted *β* values under 4X

Given our assumption for this modeling exercise that deaths are primarily occurring in the ICU, for this analysis we assumed that the information on the reported deaths is relatively accurate; hence, we do not amplify the reported deaths when fitting the death rate *υ*. Figure 10 shows the cumulative number of deaths that the model predicts under a 4X loading scenario. Note that under the 1.67X loading scenario, the ICU death rate produced by the model fit procedure was on the order of 1.24. That is, the assumption of 88% detection rate was not aligned with the point estimates we used in the model to predict the reported death rates. Given that there is widespread belief that COVID-19 deaths are underreported, we understand this finding to be in support of the idea that only a fraction of the infections are detected, and thus reported in the official case counts. In the next section, we present projections for 1.67X, 4X, and 6X loading scenarios to provide a range of future projections for cases, hospitalizations, and deaths.

**Fig 10.**
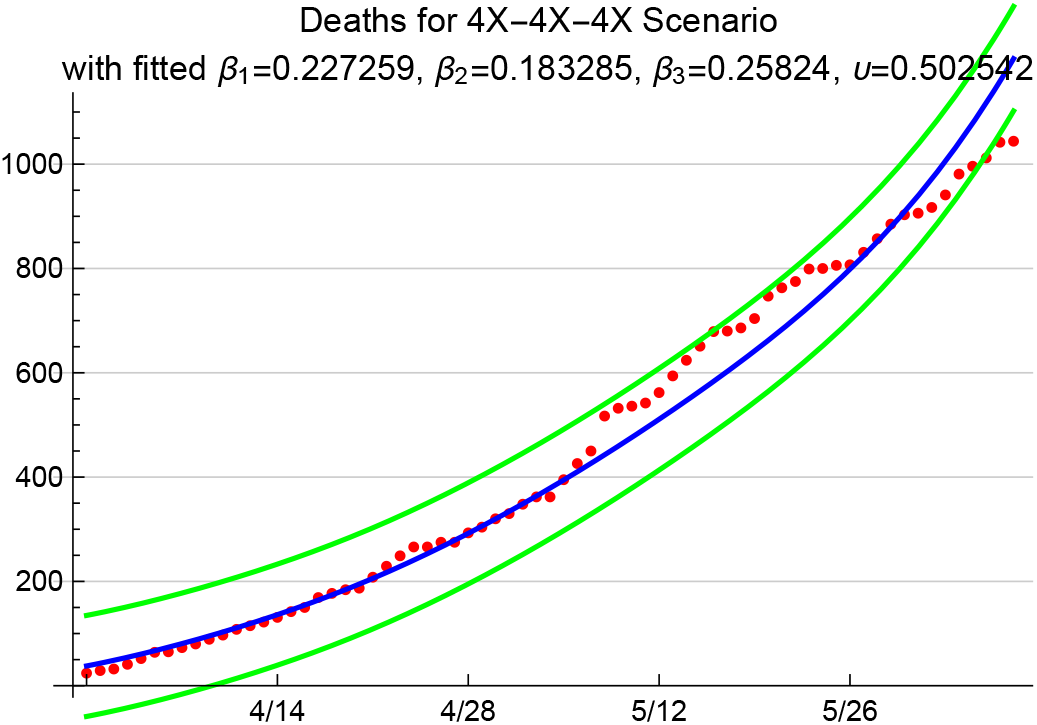
Model predicted cumulative number of deaths between 3/31 and 6/7 with 95% prediction intervals; red dots are the reported COVID-19 deaths for the period in Arizona.

## 2 Results

We provide projections on the number of deaths, number of people hospitalized, and total infections for a number of cases that differ in X-factor and the transmission rate over time. We first start with the benchmark cases of 1.67X, 4X and 6X loading scenarios simulated under the assumption that the transmission rate stays at the fitted 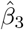 for exposure dates beyond May 24. It is useful to observe the dynamics for a relatively long horizon of 500 days, as given in Figure 11.

**Fig 11.**
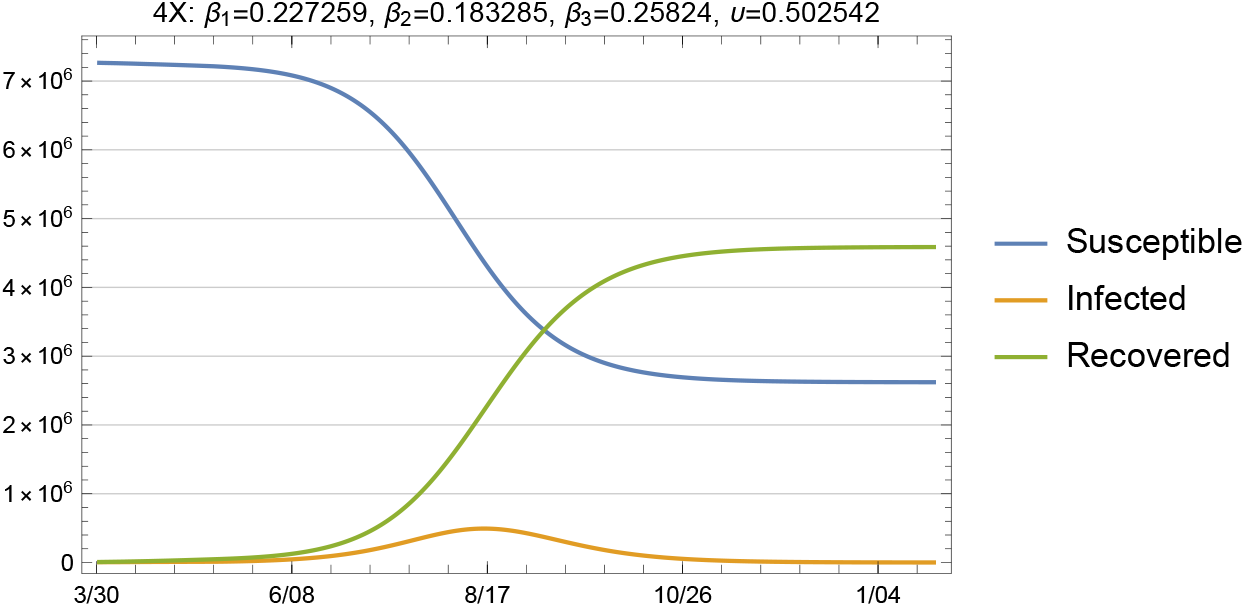
Susceptible, infected and recovered, 4X loading with fitted *β* and *υ*

Day 1 in this simulation is March 31, where we initialize our model and run it with the *β* and *υ* values that we fitted using the X-factored data on new cases and deaths after this point (i.e., 55 points of backcasted presumed exposures and 69 days of data on deaths). We define herd immunity as the point at which wide community spread is suppressed due to the large proportion of individuals in the population with immunity. Given the large initial susceptible population that we use for the model (i.e., 7,278,717) herd immunity is reached around late October at the currently fitted transmission rate. This figure demonstrates that policies that were initially enacted in March to limit close person-to-person contacts were effective in reducing transmission during April and early May, but also preserved a high pool of susceptibles in the general public to fuel future outbreaks under conditions where NPIs are not effectively implemented.

While visualization of the epi curves is useful to gain insights into the long-term behavior and other concerns such as peaks and herd immunity, it is more informative to focus on shorter-term projections since it is hard to imagine a real-life scenario under which *β* stays constant over a very long period of time due to fluctuations with regard to NPI measures taken by individuals and public health officials.

The baseline plots given in Figures 12 through 15 show the total infected and hospitalizations as well as exposures and deaths under the 1.67X, 4X and 6X loading scenarios with fitted *β* and *υ*. The fitted values in each scenario are shown in the plot legends. Recall that to fit the mortality rate in each scenario, we kept the data on reported deaths intact and fitted the value of *υ* to the data. Note that the fitted *υ* value of 0.50 for the 4X case results in a mortality rate of 2.5% among symptomatic individuals. For the 1.67X scenario, this resulted in a fitted *υ* value of 1.23, meaning that the 1.67X scenario did not generate sufficient number of patients in the ICU to explain the reported deaths in Arizona. Hence, we used an *υ* value of 0.99 for the 1.67X runs. On the other hand, the fitted upsilon for the 6X scenario was 0.33 since this scenario assumed a higher undetected rate and loaded more patients at the ICU. An alternative approach would have been to inflate the death numbers to account for the observation that deaths related to COVID-19 may be underreported. We do not use this approach in our projections in order to maintain an evidence-based conservative set of estimates on the death toll of the epidemic.

**Fig 12.**
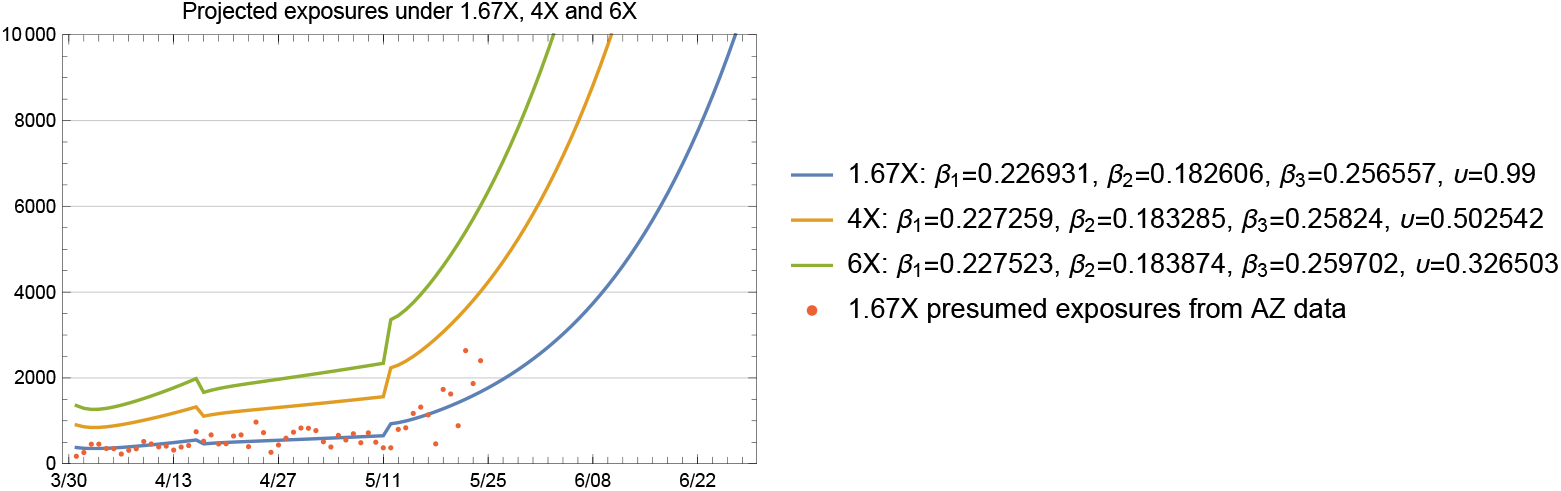
1.67X exposures inferred from actual data (red dots) and projected by the model

**Fig 13.**
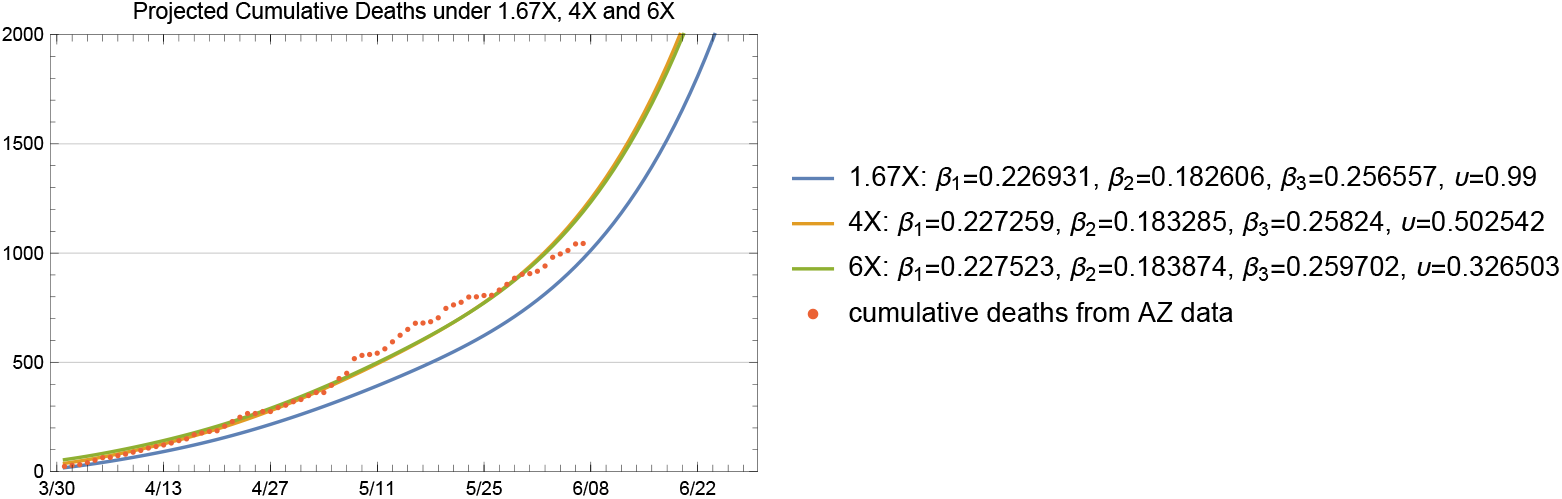
Cumulative number of deaths; actual data (red dots) and projected by the model

**Fig 14.**
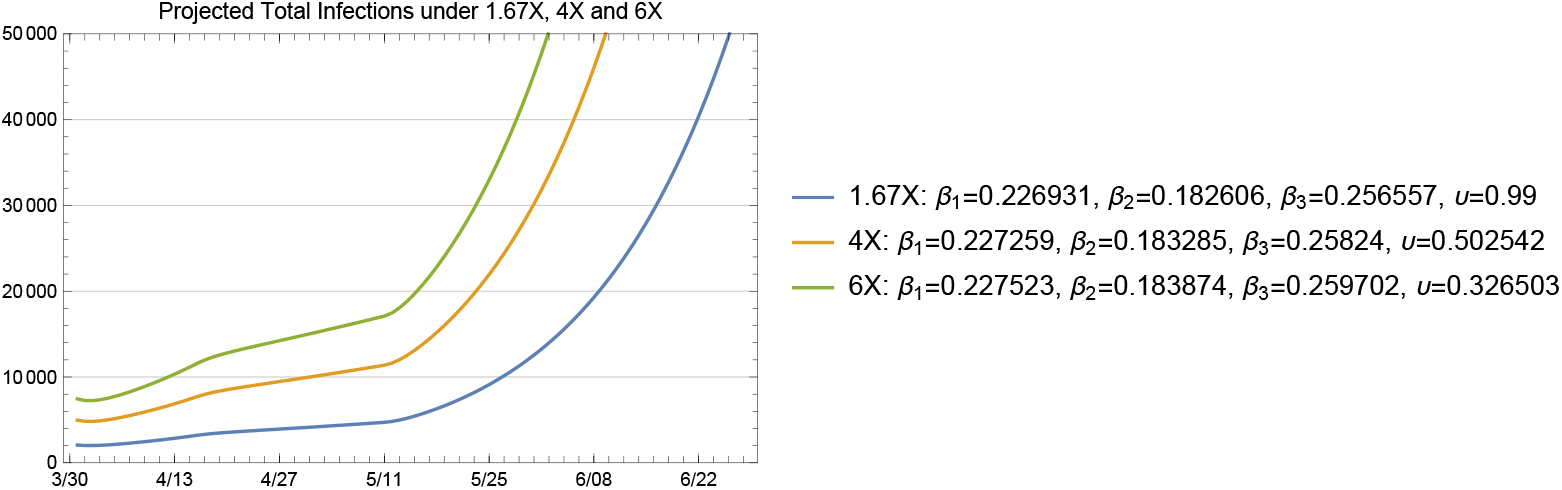
Total infected projected by the model with fitted *β* and *υ*

**Fig 15.**
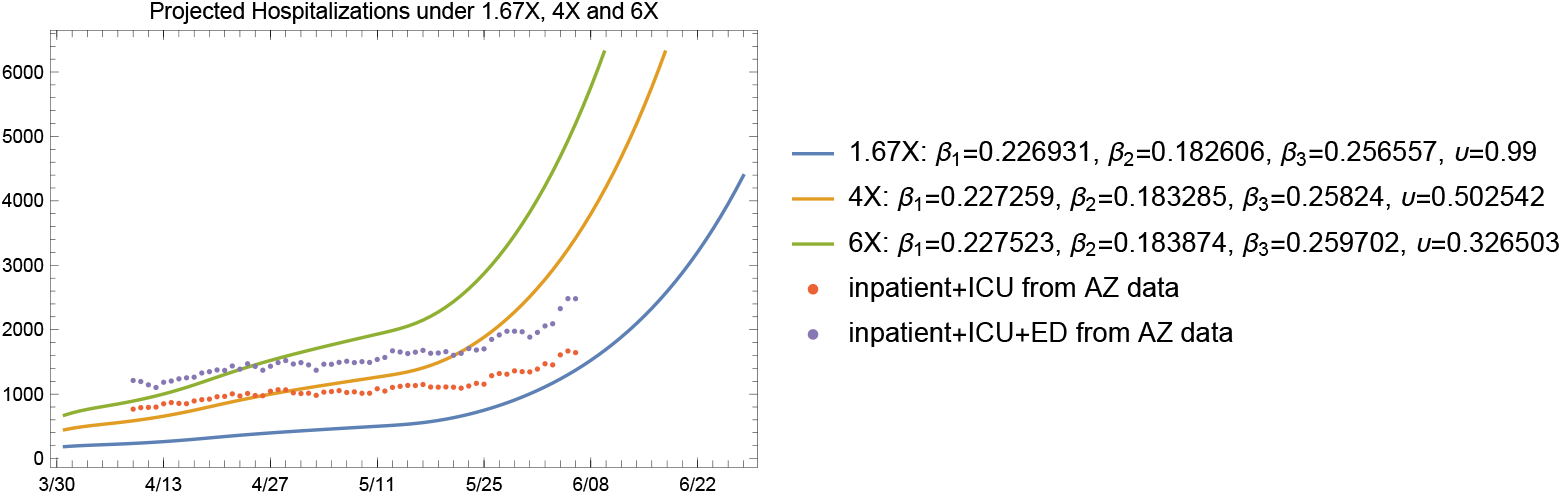
Hospitalized patients projected by the model with fitted *β* and *υ*

Figure 15 shows the projected hospitalizations (the sum of the numbers in bins 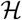, 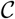, and 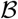) along with the available hospital data between April 9 and June 7. Again, these data are obtained from the ADHS data dashboard, where the census of inpatient, ICU, and emergency department (ED) bed usage are provided separately. Note that in our model, we do not have a separate ED bin, so we plotted the hospital data from the ADHS website in two ways: one without the numbers from the ED and one with the numbers from the ED. We note that a significant portion of the patients in the ED on each day may be discharged and sent home to recover rather than being admitted to an inpatient bed. As shown in Figure 15 the projected hospitalizations fall within the 1.67X and 4X scenarios in both of our treatments of ED data.

The model projections show that it is reasonable to expect a slowly increasing number of patients in the hospital in the short term (i.e. late May to mid June) with subsequent rapid growth of hospitalization rates with increased community transmission as NPI policies were lifted on May 15. This increase in cases is due to the large number of susceptible individuals in the population resulting, in part, to the effectiveness of NPI policies in place from March 30 to May 15.

### 2.1 Projections under a Favorable Summer Effect

There is currently debate about the impact of higher summer temperatures in large regions of Arizona on the transmission rate of COVID-19. A so-called summer effect would include a potentially suppressive effect on virus survivability in the extremely elevated temperatures and UV radiation of the desert Southwest. Simultaneously, we consider that the extreme summer heat in Arizona’s population centers also creates a behavioral effect, changing patterns of indoor and outdoor activity in Arizona’s desert environment. In essence, Arizona’s summer effect behaviorally mimics the winter effect in more temperate regions, as people seek heat relief in indoor environments. Given the uncertainty about a potential summer effect, particularly on virus survivability, we have found it to be a useful context to demonstrate the sensitivity of outbreak dynamics to the transmission rate *β*.

In Figure 16, we plotted the projected 1.67X hospitalizations under four different scenarios with respect to the summer effect. We plotted the projected hospitalizations for a longer horizon to show the impact of a favorable summer effect on transmission rates. We chose to use the 1.67X scenario for this purpose since the current hospitalization data seems to be closer to the 1.67X projections. The plot shows the tradeoff between an early summer effect versus a later but more significant summer effect. The figure also demonstrates the impact of a 25% to 50% reduction in the transmission rate as well as the impact of the timing of the summer effect in further flattening the curve.

**Fig 16.**
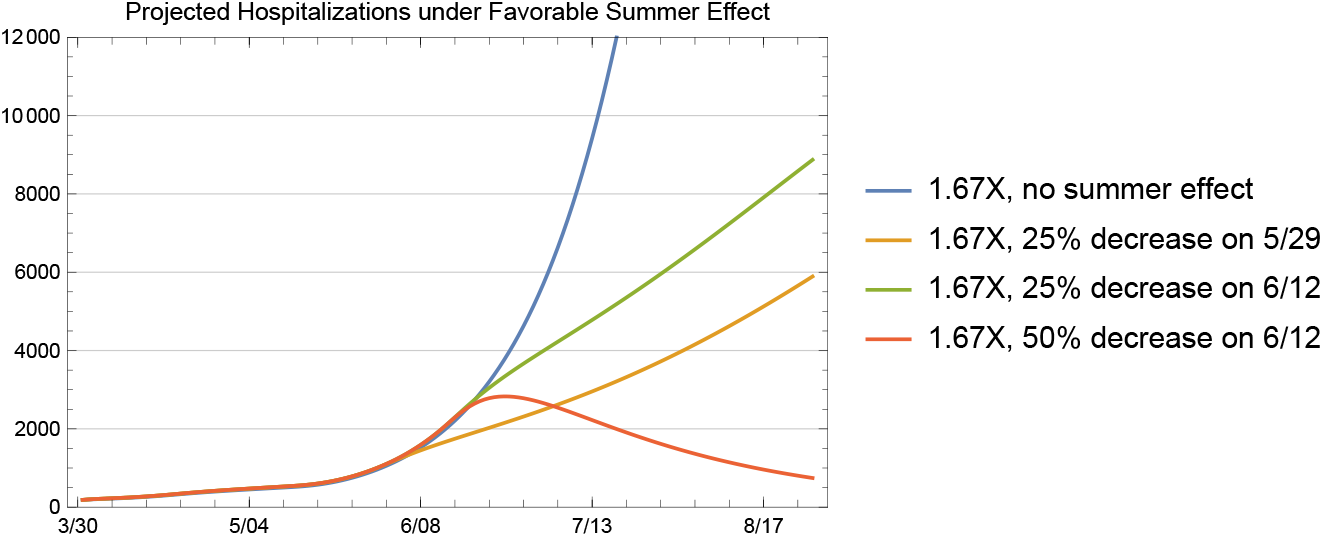
Hospitalization projections under favorable summer effect scenarios for 1.67X

### 2.2 Initiating Non-pharmaceutical Interventions

The current estimation procedure applied to the current epidemiological data as of June 7 results in a significant increase in the transmission rate starting from May 11, which we use as the transmission rate estimate to obtain baseline projections as shown in Figure 12. Of note, May 11 was the date on which restaurants were allowed to resume dine-in operations statewide. This baseline reflects the observed data from the state health department reporting system at the present time during the response. We note that there are frequent data reporting corrections and so the current estimation targets could potentially change. The model includes multiple changes in transmission rates correlating with policy implementations. When NPIs were initiated, including bar closures and restaurant restrictions in the urban centers on March 17 and the statewide stay-at-home order on March 30, transmission rates were reduced. When NPI policies were lifted, including the resumption of dine-in restaurant service on May 11 and the end of the stay-at-home order on May 15, transmission rates increased.

It is worthwhile to consider the impact of initiating NPIs once more in an effort to return to the transmission rate estimated by our model during Arizona’s stay-at-home order. We analyze the reduction in the number of infections and the number of hospitalized patients that may result from the reinitiation of NPIs at different points in time, starting from June 8. Recall that the latest data point we used in this analysis is June 7 data on cumulative confirmed COVID-19 cases and deaths, so June 8 represents the earliest point in time that the NPIs could be reinitiated. We do not specify any specific NPIs to achieve this reduction in transmission. Rather, we use the prior estimates in transmission parameter *β* as a measure of lowered transmission under heightened policy implementation.

To represent the patient care load imposed on Arizona’s hospitals, we consider the area under the total infections and hospitalizations curves, in a manner similar to the calculation of “illness inventory” or utilization in the system over time. We consider five dates that NPIs can be initiated: June 8, June 15, June 22, June 29, and July 6 and obtain the following improvement metric in comparison to the baseline case of no NPIs, which assumes that the transmission rate stays constant at the 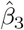. The results are qualitatively similar, so to provide some conservative estimates, we use 1.67X for this analysis. Hence, we use 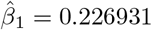, 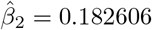, 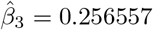. At the indicated NPI initiation times, we revert the transition rate to the lowest under NPI, which is 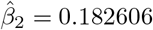.

First, it is useful to observe the total infections (including asymptomatic individuals, presymptomatic individuals, symptomatic individuals and hospitalized patients) under the baseline and the five intervention time options over the next several months under these assumptions to compare the behavior under these different scenarios. We note that we are not presenting this figure to provide projections, but rather provide a visual reference to explain the difference in long-run behavior that NPIs, through the reductions in transmission rate implies. In Figure 17 we see that the NPIs result in significantly different infection patterns as a result of the reduction in transmission rates. While this simulation represents a highly optimistic scenario with respect to the impact of NPI reinitiation on the viral transmission rate, this analysis clearly shows that initiation of NPIs can provide significant relief on the healthcare resource demands that the pandemic presents, even in the setting of elevated baseline infection rates with a large susceptible population.

**Fig 17.**
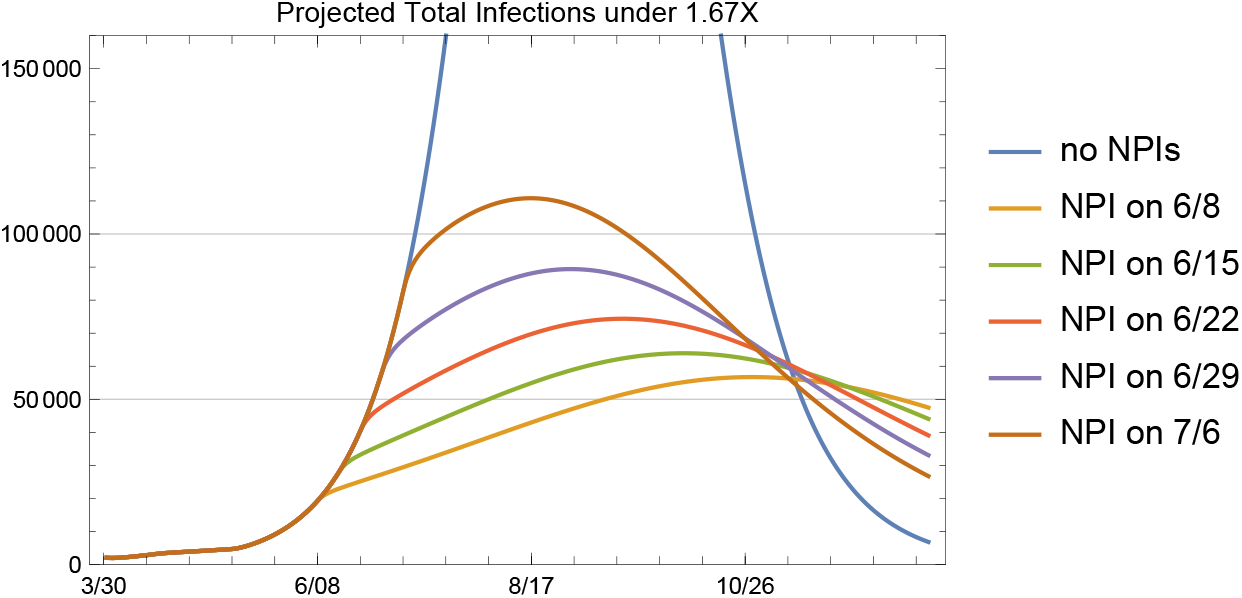
Total infections under the baseline and five NPI initiation date options for 1.67X

We have indicated that this analysis presents an optimistic scenario with respect to the impact of the re-initiation of NPIs on the transmission rate due to several reasons. First, individual behavior with regard to NPIs including masking and physical distancing varies due to individual beliefs and adherence to recommended actions. Second, some population segments experience systemic inequity including inadequate access to supply chain and economic resources to acquire protective equipment (e.g. face masks) and hygiene resources (e.g. hand sanitizer) to afford measures of protection. Third, individuals experiencing systemic resource inequities are least able to avoid public contact and practice physical distancing due to fragile employment status in front-line jobs. Fourth, the high asymptomatic rate and presymptomatic transmission patterns coupled with low viral testing rates and extremely limited contact tracing capacity statewide has limited the ability of the public health system to contain outbreaks even in the setting of optimal NPI adherence by individuals in the community.

To quantify the improvement that can be expected from the re-initiation of NPIs on a given date, we consider the following percent reduction metric, which compares the areas under each curve over time. That is,

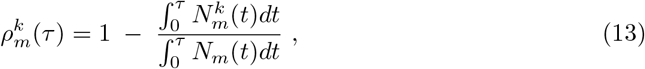

where the superscript *k* denotes the NPI initiation date options and superscript *m* can be total infections, hospitalized patients or deaths. We calculate the reduction metrics for 1.67X however, the results are similar qualitatively under 4X and 6X.

Table 2, which provides the percent reductions in total infections, shows that initiating NPIs results in significant reductions in total infections. For example, initiating NPIs on June 22 would imply a reduction of 72% in the total infections in Arizona by the beginning of September. A similar observation can be made from Table 3; initiation of NPIs on June 22 would also imply 68% reduction in hospitalizations.

**Table 2.** Percent reductions observed in the total infections by the indicated dates for each NPI initiation date option

| Percent reduction in total infections with NPI initiations |  |  |  |  |  |
| --- | --- | --- | --- | --- | --- |
| by date | NPI on 6/8 | NPI on 6/15 | NPI on 6/22 | NPI on 6/29 | NPI on 7/6 |
| 7/1/20 | 24% | 13% | 4% | 0% | 0% |
| 8/1/20 | 65% | 57% | 48% | 38% | 27% |
| 9/1/20 | 82% | 77% | 72% | 65% | 58% |
| 10/1/20 | 84% | 80% | 76% | 71% | 65% |
| 11/1/20 | 82% | 78% | 74% | 69% | 64% |
| 12/1/20 | 78% | 74% | 70% | 66% | 61% |

**Table 3.** Percent reductions observed in the hospitalizations by the indicated dates for each NPI initiation date option

| Percent reduction in hospitalizations with NPI initiations |  |  |  |  |  |
| --- | --- | --- | --- | --- | --- |
| by date | NPI on 6/8 | NPI on 6/15 | NPI on 6/22 | NPI on 6/29 | NPI on 7/6 |
| 7/1/20 | 14% | 5% | 1% | 0% | 0% |
| 8/1/20 | 58% | 49% | 39% | 28% | 17% |
| 9/1/20 | 79% | 74% | 68% | 61% | 52% |
| 10/1/20 | 84% | 80% | 75% | 70% | 64% |
| 11/1/20 | 83% | 79% | 74% | 70% | 64% |
| 12/1/20 | 79% | 75% | 71% | 67% | 62% |

Another important insight we gain from the results is the impact that timing of the NPIs makes. In both tables, we see that the reductions by July 1 for the initiation dates of June 29 and July 6 are 0%, since those cases have the same behavior as the baseline 1.67X case. In general, supposing that June 8 would be the earliest that one would trigger an NPI, delaying the initiation of NPIs by one week results in about a 5% change in the reductions that we observe in the total infections, hospitalizations and deaths. This assessment model may provide an estimate for the public health burden of each week of delaying policy enactment or individual practice of NPIs.

Another percent reduction metric that one could look at is the maximum number of patients hospitalized under each case. Since the start of the epidemic, the peak hospital resources required to care for COVID-19 patients has been an important concern among public officials indicating the need to flatten the curve. Initiating NPIs on June 8, June 15, June 22, June 29 and July 6 result in respectively 88%, 86%, 84%, 81% and 77% reductions in the maximum number of patients hospitalized (i.e. the peak of each curve). Considering the limitations in the hospital and particularly ICU resources, including expert healthcare personnel, to provide safe and effective care for seriously ill COVID-19 patients as well as patients with critical conditions unrelated to COVID-19, we note that these resource utilization reductions resulting from NPI policy enactment may make a significant difference in population health outcomes.

In addition to the percent reductions in the areas under the total infections and hospitalized curves, we present in Table 4 the percent reductions in the number of deaths by the dates indicated in the first column. Even without considering any negative effects of exceeding hospital care capacity (which is likely to happen without the initiation and widespread adoption of NPIs) on patient health outcomes, we see that reductions in deaths resulting from the initiation of NPIs is on the order of 70%. Again, our model demonstrates that one week’s delay in the initiation of NPIs corresponds to a 4% to 10% difference in the reductions in deaths.

**Table 4.** Percent reductions in deaths by the indicated dates for each NPI initiation date option

| Percent reduction in deaths with NPI initiations |  |  |  |  |  |
| --- | --- | --- | --- | --- | --- |
| by date | NPI on 6/8 | NPI on 6/15 | NPI on 6/22 | NPI on 6/29 | NPI on 7/6 |
| 7/1/20 | 10% | 4% | 0% | 0% | 0% |
| 8/1/20 | 54% | 44% | 34% | 23% | 14% |
| 9/1/20 | 78% | 72% | 66% | 58% | 49% |
| 10/1/20 | 84% | 80% | 75% | 69% | 63% |
| 11/1/20 | 83% | 79% | 75% | 70% | 65% |
| 12/1/20 | 80% | 76% | 72% | 67% | 62% |

## 3 Discussion

In this paper we have proposed a methodology for modelling and projecting the spread of the COVID-19 epidemic in Arizona by considering publicly available data from March 4 (first date with a confirmed COVID-19 case with community spread in Arizona) to June 7. This work is focused on understanding features of infection and disease transmission, as well as exploring the impacts of possible scenarios for implementing control measures through public policy. We note that this model iteration was initially constructed beginning in April at the time of a statewide stay-at-home order, and refined after the stay-at-home order was lifted. This timeframe of the model iteration process allowed for clear observation of the dynamic transmission rates in response to public policy implementation and individual adoption of NPI behaviors.

There are several limitations to our analysis. It is important to note that our SEIRD modeling approach did not take into account many factors that play an important role in the dynamics of disease such as heterogeneous contact transmission network, the characteristics of the population (e.g. age, comorbid health conditions, racial and ethnic disparities in access to testing and treatment), the possibility of partial immunity or no immunity from SARS-CoV-2 infection and the availability of testing and contact tracing. At the time of this report, Arizona maintained one of the lowest per capita testing and contact tracing rates of any state in the country. Therefore, it is likely that significant underdetection and thus underreporting of mild and asymptomatic cases may impact calculations of hospitalization and death rates. In order to accommodate this limitation, we used biologically plausible parameters for SARS-CoV-2 based on current evidence. As the evidence related to SARS-CoV-2 and COVID-19 continues to develop, these values are likely to be updated as more comprehensive data become available.

In future work, we look forward to testing this model more broadly against data from other states beyond Arizona in an effort to validate this approach for other public health policy making jurisdictions. In addition, we plan to test this modeling approach more narrowly by applying it to county-specific data in Arizona in order to assess the retrospective accuracy given a more homogeneous population sample of a single county as opposed to the highly heterogeneous population sample represented by the full state of Arizona. We anticipate that this type of comparative work may inform best practices for projection modeling in future epidemic conditions. Establishing best practices for projection modeling can, in turn, provide improved inputs for policymakers with more clear expectations and understanding about the scope and limitations of models in highly uncertain conditions like the current COVID-19 pandemic.

Our work illustrates how a locally contextualized system dynamics model can be very useful for making inferences about how the pandemic impacts may change in response to policy and individual behavior decisions about implementation of different disease mitigation measures.

## Data Availability

All data used is publicly available on https://covidtracking.com/ and the https://www.azdhs.gov/preparedness/epidemiology-disease-control/infectious-disease-epidemiology/covid-19/dashboards/index.php.

https://biodesign.asu.edu/research/clinical-testing/critical-covid-19-trends

## Acknowledgments

We would like to acknowledge the contributions of Heidi Gracie and Joshua LaBaer, and the Arizona Department of Health Services Modeling Working Group including Amber Asburry, Steven Robert Bailey, Timothy Flood, Joe Gerald, Katherine Hiller, Ken Komatsu, Mark Manfredo, Vern Pilling, Timothy Richards, Marguerite Sagna, Lisa Villarroel, Patrick Wightman, and Neal Woodbury.

